# In-Situ Disinfection of Hospital Privacy Curtains Using a Next-Generation Aerosolized Hydrogen Peroxide System

**DOI:** 10.64898/2026.10.21.26365806

**Authors:** Cristofer Navas, Barbara Hawkins, Catherine Lazarus, Erik Kulstad, Kathy Warye

**Affiliations:** Mel & Enid Zuckerman College of Public Health, University of Arizona, Tucson, AZ, United States of America; Little Colorado Medical Center, Winslow, AZ, United States of America; Northwestern University, Evanston, IL, United States of America; University of Texas Southwestern Medical Center, 5323 Harry Hines Blvd, Dallas, TX, United States of America; Infection Prevention Partners, San Francisco, CA, United States of America

## Abstract

**Background:** Privacy curtains represent a uniquely difficult infection control surface because they are frequently touched by patients, visitors, and healthcare workers, yet are difficult to disinfect using routine environmental cleaning protocols. We sought to quantify the performance of a novel automated aerosolized hydrogen peroxide system against pathogens on privacy curtains.

**Methods:** This study was performed in an enclosed office space (≈ 65 m³), approximating a patient exam room. Four *Bacillus atrophaeus* (ATCC 9372) biological indicators (BIs) were affixed to a standard polyester privacy curtain. A single one-minute fogging phase was followed by a ten-minute contact/aeration phase (total = 11 min). Four corresponding chemical indicators measured peak airborne aerosolized hydrogen peroxide concentration, and two control BIs were placed outside of the room.

**Results:** All four BIs demonstrated complete inactivation (no growth after seven-day incubation), representing ≥4-log_10_ spore reduction. Peak dosages reached 42 ppm nearest the aerosolized hydrogen peroxide jet outlet, with the lowest value at the near point on the back side of the curtain reaching 27 ppm (exceeding the target dosage of 25 ppm). Control exposures failed to achieve sterilization.

**Conclusions:** A next-generation aerosolized hydrogen peroxide dry-fogging disinfection system achieved full inactivation of resistant spores on both sides of privacy curtains within a short 11-minute automated cycle.

## Background

Reducing the national burden of healthcare-associated infections (HAIs) remains a core patient safety priority. With 1 in 31 U.S. patients contracting HAIs daily, it is crucial to address the gap in HAI research and literature.^1^ Further data collection on pathogen reservoirs, transmission, and HAI prevention is pivotal to informing the disinfection policies and protocols implemented across diverse healthcare facilities. High-touch surfaces, such as privacy curtains, are frequently contaminated with healthcare-associated pathogens and can serve as reservoirs for cross-transmission, with microbial contamination found in 42–92% of curtains within three weeks of placement in clinical environments.^2^ Unlike hard nonporous surfaces, textiles such as privacy curtains present a complex disinfection target due to fiber depth, folds, and occluded surfaces that can limit direct exposure to conventional disinfectants.

The Centers for Disease Control and Prevention (CDC) recommends that facilities remove and launder hospital curtains after a patient discharge or transfer, with a recommended frequency of monthly, if not visibly soiled.^3^ However, current disinfection policies and procedures remain variable between healthcare facilities. Traditional laundering of curtains is labor-intensive and cannot feasibly be performed at the frequency required to prevent recontamination.

Previous studies have proven that the use of hydrogen peroxide in vapor, liquid, and aerosol forms is effective in reducing pathogen presence on hospital curtains.^4,5^ This method serves as an alternative to the challenges presented by ultraviolet (UV)-C light disinfection. The efficacy of UV-C disinfection, while widely used and accepted, is limited by difficult topography.^6^ In contrast, aerosolized disinfection may provide more uniform exposure across complex curtain geometries. Thus, the goal of this study was to investigate the efficacy of automated disinfection with a next-generation aerosolized hydrogen peroxide dry fogging system, which offers a potential in-situ alternative capable of achieving effective deposition across exposed textile surfaces, including folded or partially shielded regions of fabric surfaces and hard-to-reach areas.

## Methods

### Study design and setting

This pre-clinical validation study was performed in an enclosed office space approximating a patient room or an operating room suite (Figure 1).

**Figure 1.**
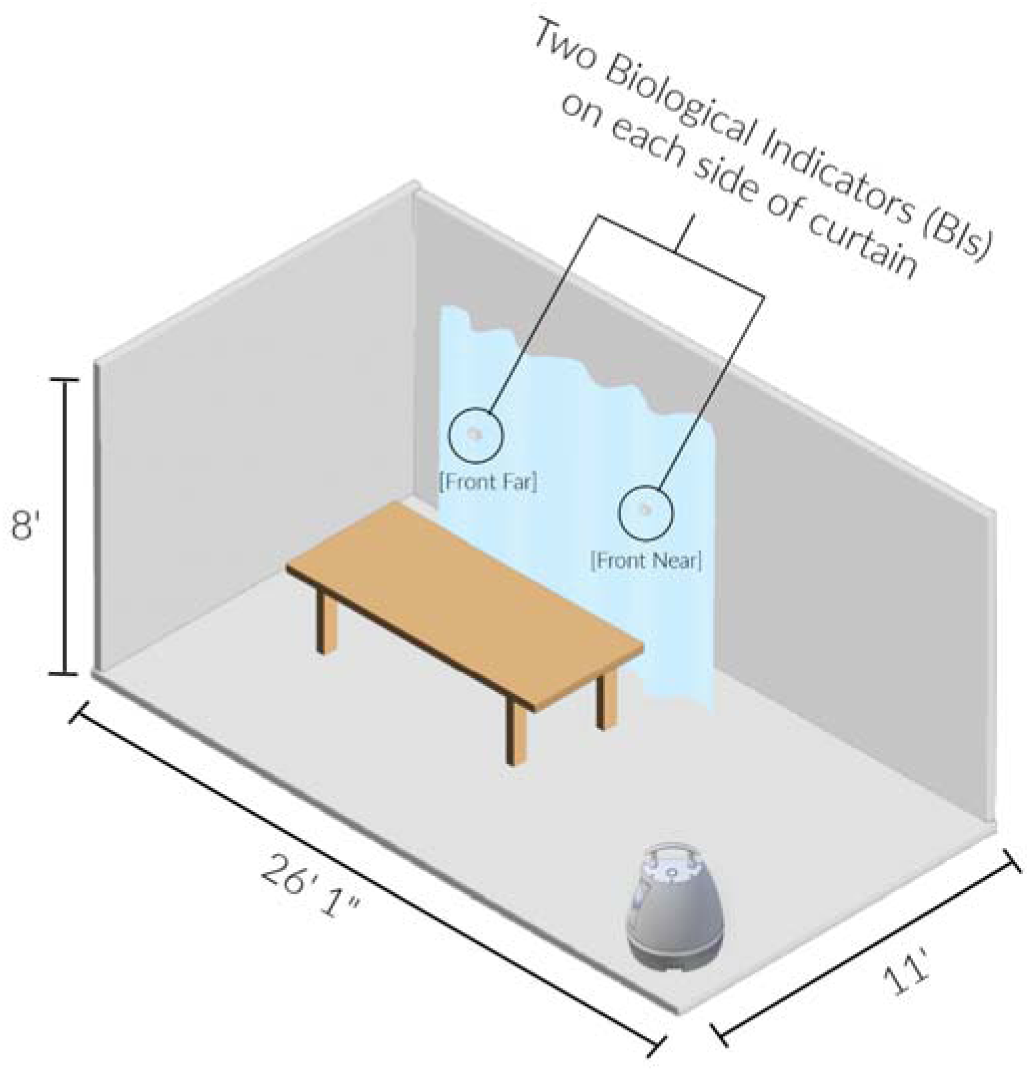
Diagram of Room Configuration and Placement of Automated Aerosolized Hydrogen Peroxide Dry Fogging System. Specifically, room dimensions were 11 ft by 26 ft with an 8 ft ceiling (for a total volume of 2288 cubic feet, or 64.8 cubic meters). The space included a desk, chairs, carpeting, a computer, and various boxes throughout (Figure 2).

**Figure 2.**
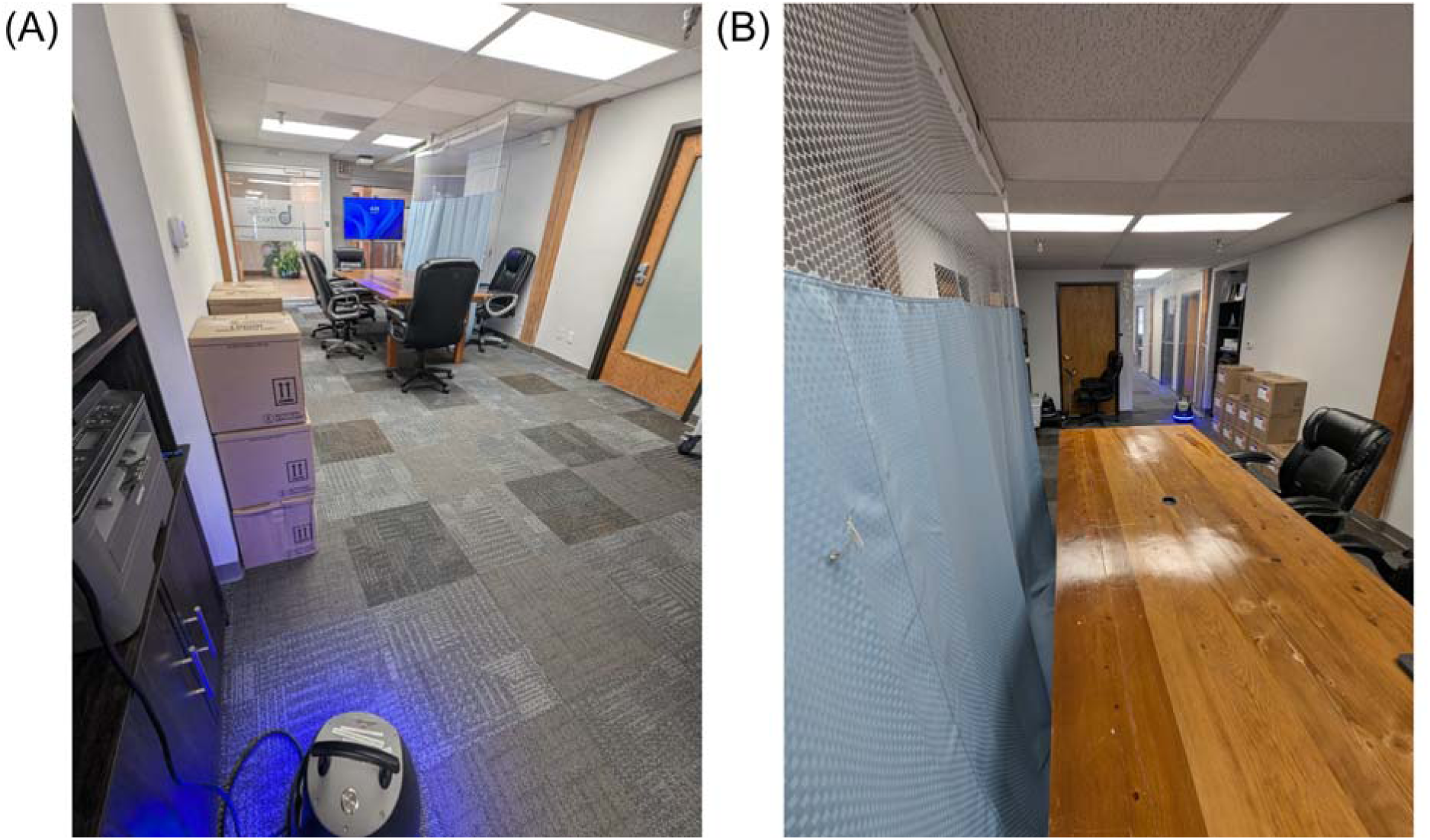
Test Room Setup. (A) View of the testing room from the side nearest the automated aerosolized hydrogen peroxide dry fogging system and (B) from the side farthest from the automated aerosolized hydrogen peroxide dry fogging system.

The device used for this study was an automated aerosolized hydrogen peroxide dry fogging system (Breezy Med, Albuquerque, NM, USA).^7^ This novel technology (Figure 3) provides advanced aerosolization via high-flow jets (4.09 m³/min airflow) to provide micron-sized hydrogen peroxide droplets that disperse throughout the entire room, ensuring optimal coverage of surfaces.

**Figure 3.**
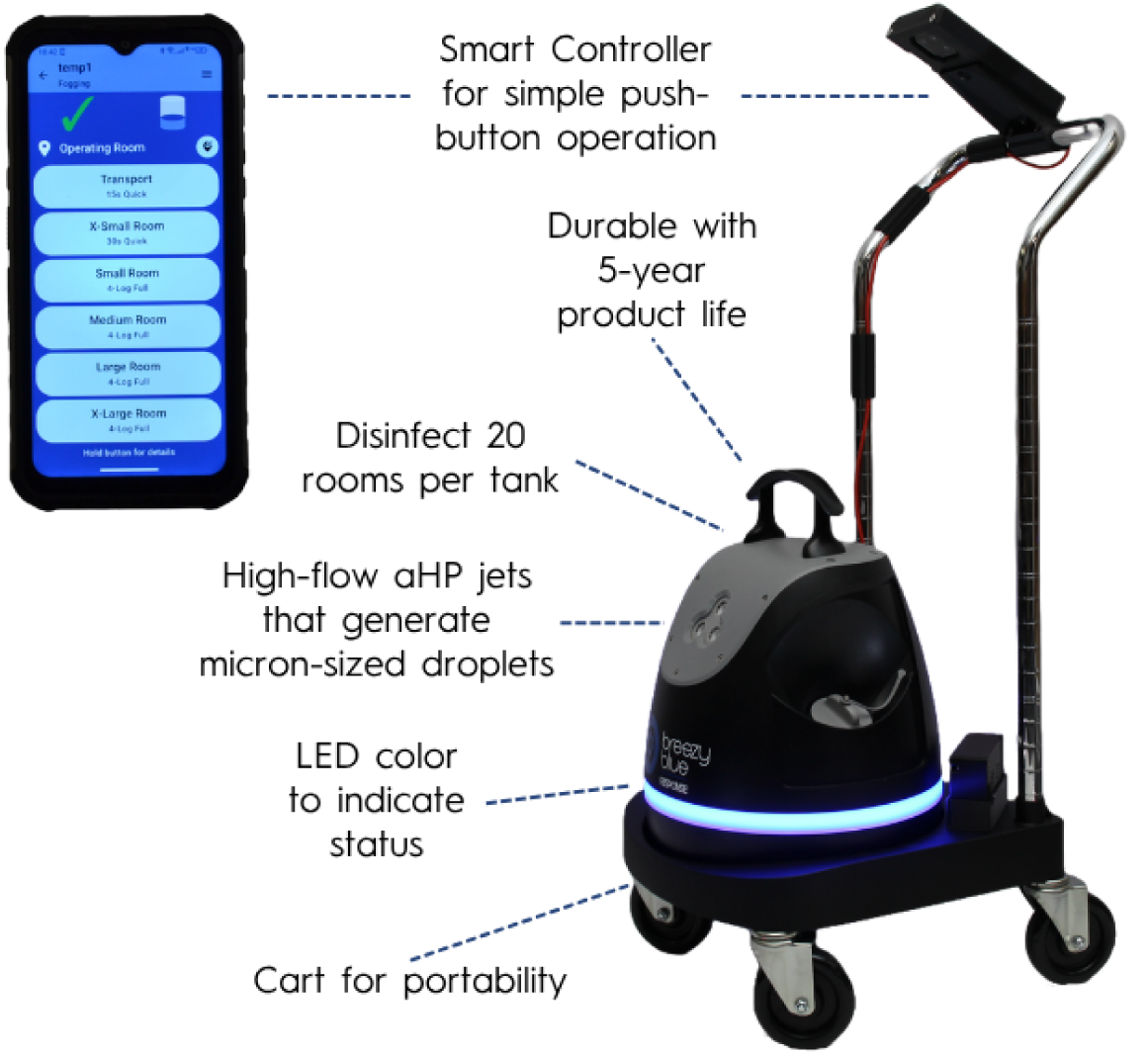
The Next-Generation Automated Aerosolized Hydrogen Peroxide Dry Fogging System. (Breezy Med, Albuquerque, NM, USA).

The micron-sized droplets are intended to remain suspended within the enclosed treatment space, minimizing escape beyond closed doors and windows. The system also allows integration within data cloud platforms to provide historical data on equipment usage, verification of aerosolized hydrogen peroxide dosages, room coverage, disinfection times, and locations for quality assurance and auditable records.

### Experimental procedure

The hydrogen-peroxide formulation used in the device contained 5.0% H□O□ with 0.01% silver ions. Four *Bacillus atrophaeus* (ATCC 9372) biological indicators (BIs) with >10□ spores each (Lioi Chem BI’s and Broth Kit PN: 91065 (4-LOG) or 91064 (5-LOG), Roseto degli Abruzzi, Italy) were affixed with pins to a standard polyester privacy curtain (Figure 4).

**Figure 4.**
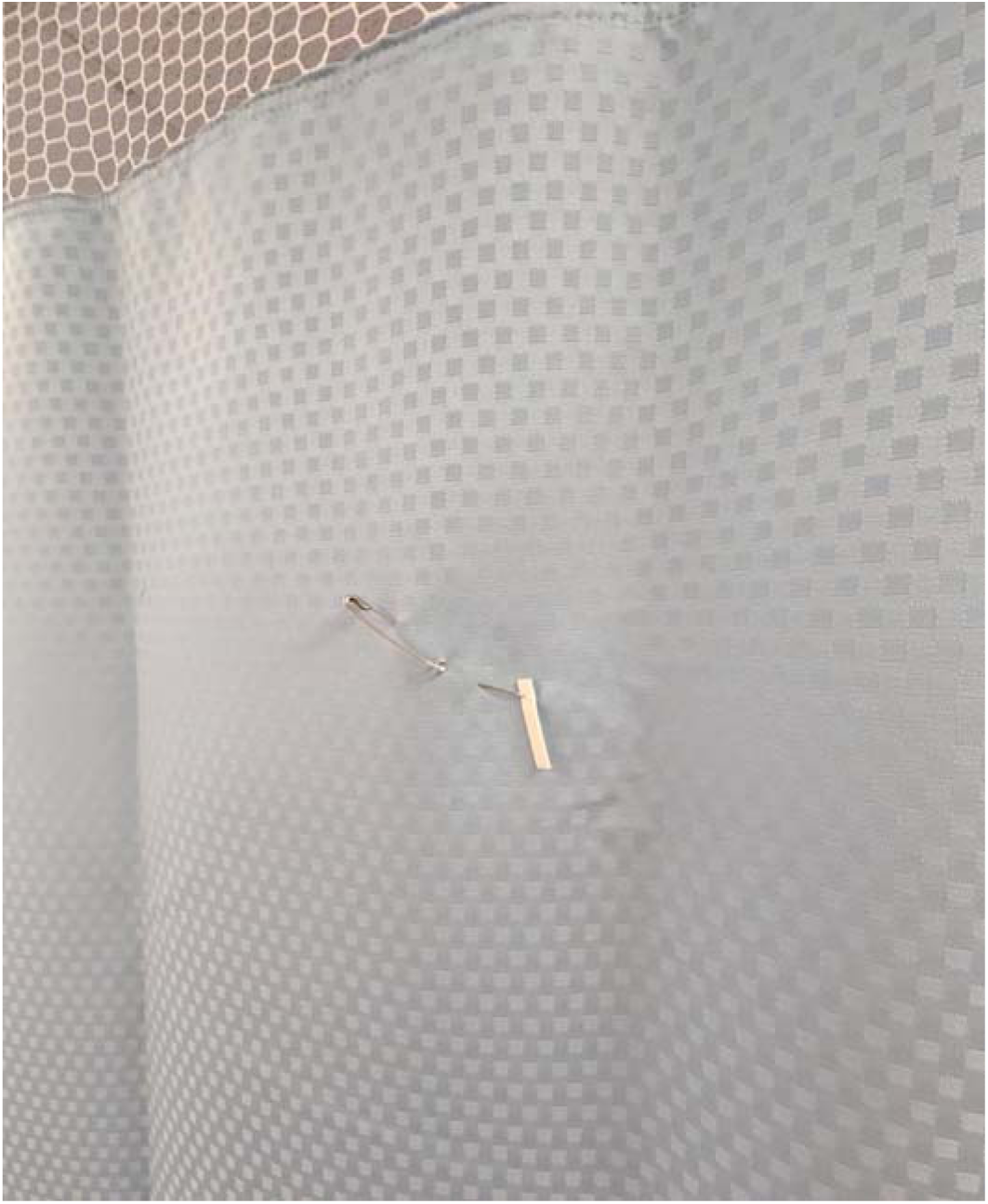
Placement of Biological Indicator (BI) on the Privacy Curtain. The BI containing Bacillus atrophaeus (ATCC 9372) with >10⁴ spores each affixed with a pin to a standard polyester privacy curtain. A total of 4 BIs were placed on the curtain at the front and back surfaces and at near and far positions relative to the aerosolized hydrogen peroxide jet outlet.

*Bacillus atrophies* spores were selected as a more strenuous biological challenge because spore-forming organisms exhibit substantially greater resistance to disinfection than vegetative bacteria and many enveloped viruses, serving as useful biological indicators for sterilization.^8^ Locations for the BIs were on the front and back surfaces of the curtain at near and far positions relative to the aerosolized hydrogen peroxide jet outlet (as shown in Figure 1). Four corresponding chemical indicators (Breezy Trace, an electrochemical sensor system measuring 0 to 100 ppm range with 10% accuracy) were used to measure peak airborne concentration. A single one-minute fogging phase was followed by a ten-minute contact/aeration phase, for a total treatment time of 11 minutes. Following the treatment, the BIs were immediately removed, placed into the growth medium, and incubated for 7 days at 32.5° C.

### Study measures

Hydrogen peroxide concentration was measured and recorded across the treatment time, as manufacturer data suggest a minimum threshold of 25 ppm hydrogen peroxide concentration to achieve a 4-log reduction. Biological indicators were incubated for 7 days at 32.5° C to determine the presence of *Bacillus atrophies’* growth. Two control BIs were placed outside of the room and, therefore, were not exposed to hydrogen peroxide, to confirm the viability of the BIs.

## Results

### Biological indicators

After the total cycle time, including fogging, contact, and passive aeration, of 11 minutes, all four BIs demonstrated complete inactivation (no growth after seven-day incubation), representing ≥4-log_10_ (≥ 99.99%) spore reduction. Control BIs exhibited expected growth after 2 days of incubation, confirming activity of the test system.

### Hydrogen peroxide concentration

Peak hydrogen peroxide concentrations reached as high as 42 ppm (Figure 5) nearest the aerosolized hydrogen peroxide jet outlet, with the lowest value of 27 ppm occurring at the near point on the back side of the curtain (exceeding the target dosage of 25 ppm).

**Figure 5.**
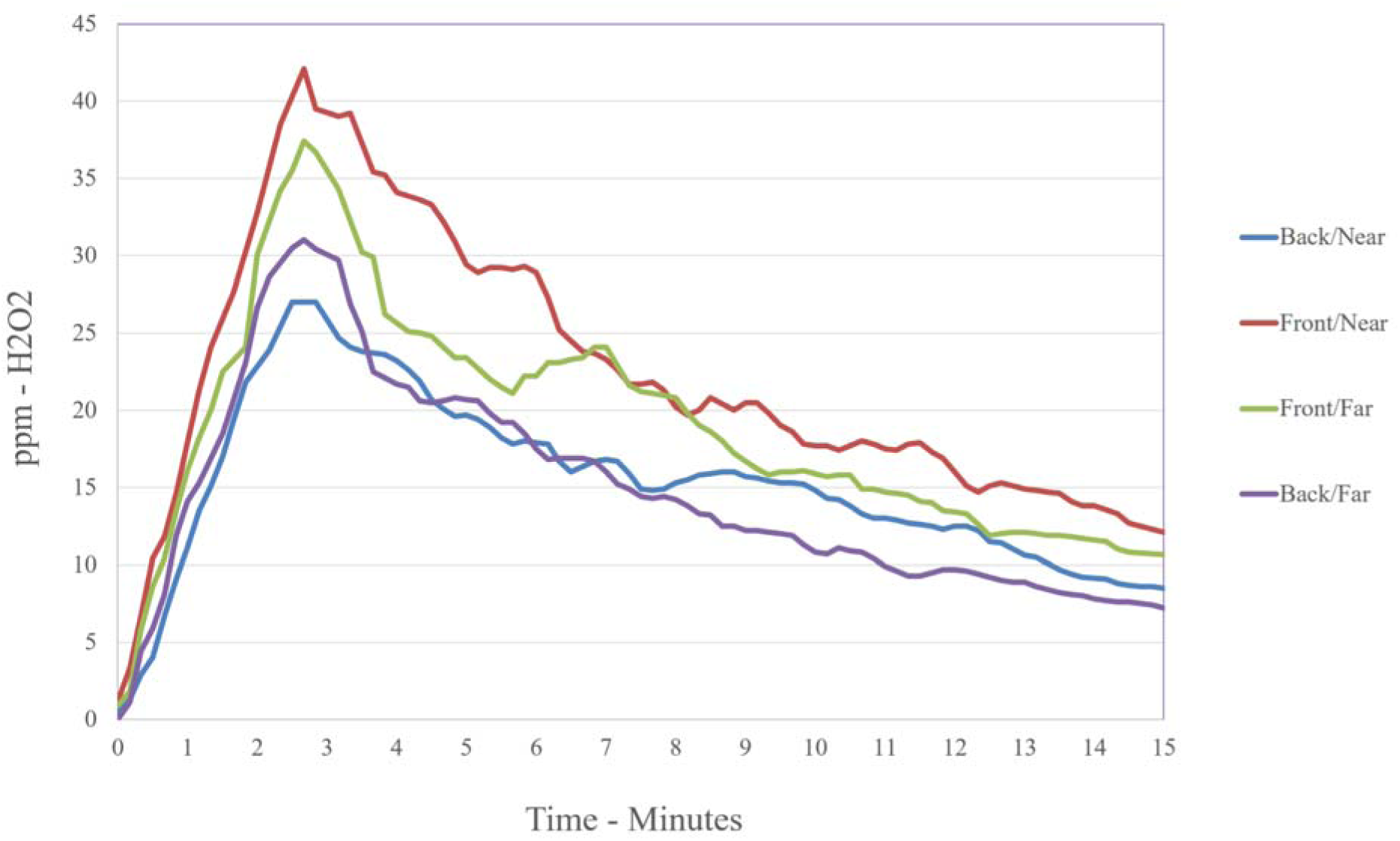
Hydrogen Peroxide Concentration During Treatment Time for Each Monitored Site. Peak dosages reached as high as 42 ppm nearest the aerosolized hydrogen peroxide jet outlet, with the farthest point on the back side of the curtain reaching 27 ppm (exceeding the target dosage of 25 ppm).

Peaks for each location specifically were 42 ppm for the front/near site, 37 ppm for the front/far site, 27 ppm for the back/near site, and 31 ppm for the back/far site.

## Discussion

In this pre-clinical validation study, a next-generation aerosolized hydrogen peroxide dry-fogging disinfection system achieved complete inactivation (≥4-log_10_ spore reduction) of resistant spores on both sides of privacy curtains within a short 11-minute automated cycle. These findings suggest that rapid in-situ aerosolized disinfection may offer a practical alternative to manual laundering or line-of-sight disinfection methods for textile barriers in patient care environments.

The challenges of curtain disinfection are significant in high-throughput clinical environments; consequently, disinfection of curtains is often deprioritized in the wake of competing operational imperatives. This is partially due to the difficulty of disinfecting curtains using customary workflows and existing methods.^9^ With privacy curtains frequently touched by healthcare workers, patients, and visitors, there are many opportunities for contamination and modes of transmission. Healthcare workers can unknowingly contaminate curtains while wearing gloves or after having touched other surfaces.^10^ In conjunction with being a high-touch surface, curtains serve as effective reservoirs for pathogens due to their highly porous and absorbent material, even if designed with antimicrobial properties.^11,12^ Curtains are also positioned in proximity to patients and subject to contamination by pathogens known to be airborne, such as Methicillin-resistant *Staphylococcus aureus* (MRSA) and *Clostridium difficile* (*C. difficile*). Current disinfection protocols rely on reactive conventional laundering of curtains when visibly soiled. This practice results in highly variable cleaning cycles, prolonged pathogen exposure and contact times, and logistical burdens on environmental services (EVS).

Alternative methods of disinfecting curtains include ultraviolet(UV)-C, steam cleaning, spray spot cleaning, and other hydrogen peroxide solutions via diverse delivery methods.^13^ Although methods such as UV-C disinfection are available, textile surfaces are particularly challenging for UV-C systems; shadowed regions that reduce photon exposure, repositioning, and time-to-result prolong clinical workflows and can yield inconsistent decontamination.^14,15^ Prior work evaluating UV-C decontamination of inoculated textiles demonstrated limited microbial reduction despite prolonged exposure times. Evaluation of an automated UV-C room disinfection device’s ability to decontaminate textiles inoculated with *Enterococcus faecium* demonstrated that decontamination reduced *E. faecium* counts by a mean log_10_ reduction factor of only 1.37 (p = 0.005) with a required median time of 111 minutes.^16^ The comparative efficacy of this method relative to aerosolized hydrogen peroxide remains subject to investigation.

Similar modalities to aerosolized hydrogen peroxide, such as plasma-treated hydrogen peroxide mist (PTHPM) disinfection, are undergoing clinical evaluation.^17^ A recent *in-situ* study conducted by Choi et al. validated the broad microbicidal advantages of hydrogen peroxide; however, while the integration of plasma treatment generates highly enhanced reactive species, it yields a transient lifespan and introduces substantial technical complexity.^18,17^ Furthermore, the study sampled various high-touch surfaces, but did not assess the efficacy of PTHPM on porous textiles, such as privacy curtains.^17^ User training, labor and storage, as well as the safety risks to humans must also be considered when employing this technology. The efficacy of aerosolized hydrogen peroxide systems depends not solely on hydrogen peroxide concentration, but also on aerosol dynamics including droplet size distribution, spatial dispersion, evaporation rate, and air mixing. Micron-scale droplets can remain suspended long enough to distribute throughout the treatment space and deposit onto complex surfaces, including folded textile materials that are difficult to access with wipe-based or line-of-sight technologies. Mathematical modeling studies have been performed on the system to efficiently visualize and quantify droplet dispersion and determine room coverage.^7^ Mathematical modeling of this process is useful, since the dry fogging is otherwise difficult to detect with the naked eye. At the velocities quantified in this analysis, room coverage from surface contact with hydrogen peroxide in a standard room size of 11 feet x 12 feet with an 8 foot ceiling was complete less than 1 minute into the treatment course.^7^ The animation accompanying the analysis by Lazarus et al. provides a visualization of the physical movement of particles, and suggests a rationale for the concentrations detected in this study.^7^ In particular, the highest concentration of hydrogen peroxide is in the expected location, closest to the jet outlet, while the lowest concentration is distal to this point, but not at the most distant point from the jet outlet. The airflow dynamics in typical room settings are such that the fog would circulate back in a pathway to the back side of the curtain, resulting in the pattern seen in this study.

Hydrogen peroxide, via various mediums, has been suggested as a treatment option for various areas of the hospital, including curtains. Cadogan et al. tested hydrogen peroxide spray and wipes on hospital privacy curtains in a regional burn/plastic surgery ward.^19^ Treatment groups were cleaned with two commercially available products: a hydrogen peroxide 1.4% disinfectant spray and a hydrogen peroxide 1.4% disinfectant wipe. Two groups were cleaned at 3–4-day intervals with either the disinfectant spray or wipe (a control group was not cleaned). By day 21, the control group was statistically more contaminated (2.2 CFU/cm^2^) than the spray group (1.3 CFU/cm^2^) or wipe group (1.5 CFU/cm^2^) (p < 0.05). After each cleaning (performed at 3–4-day intervals), the bacterial burden on the curtains reduced to near day 0 levels; however, the level was found to increase again over the intervening 3-4 days. By day 21, 64% of control curtains were contaminated with MRSA compared to 10% (spray) and 5% (wipe) (p < 0.05).^19^ Details on the time required for manual spraying or wiping were not provided. McKew et al. investigated an older hydrogen peroxide system acquired in 2014 for efficacy against MRSA, VRE, Gram-negative multi-resistant organisms and other Gram-negative bacilli on various surfaces in a burns unit.^20^ In general, they found a reduction in the rates of contaminated surfaces with this older system, with a marked reduction in MRSA contamination of soft surfaces (12% post-clean vs 6% post-aerosolized hydrogen peroxide), and patient room surfaces (8.3% post-clean vs 2.8% post-aerosolized hydrogen peroxide), but limited efficacy against MRSA in bathrooms. Although the study concludes that the older system is a useful method of environmental disinfection, the soft surfaces tested included bed mattresses, pillows, patient recliners, and visitors’ chairs, but did not include porous surfaces or curtains.^20^ Barbot et al. evaluated an older generation hydrogen peroxide dry-mist system and a 0.5% hypochlorite solution with respect to their ability to disinfect *C. difficile*-contaminated surfaces.^21^ Before disinfection, 24% of samples obtained in rooms randomized to hypochlorite treatment and 19% of samples obtained in rooms randomized to hydrogen peroxide treatment showed environmental contamination. After disinfection, 12% of samples from hypochlorite-treated rooms and 2% of samples from hydrogen peroxide-treated rooms showed environmental contamination, representing a 50% reduction in contamination after hypochlorite decontamination and 91% after hydrogen peroxide decontamination (p < 0.005). In this study, surfaces included the toilet seat, bathroom sink, bathroom floor, bedside table, care table, telephone, door handle, chair arm, floor of the main room, windowsill, remote control for the bed, bed barrier, and alcohol-based hand gel dispensers, but did not include curtains.^21^ Mosca et al. compared the effectiveness of an older system using hydrogen peroxide <8% with silver ion versus a manual method with 0.5% sodium hypochlorite solution.^22^ Disinfection with the hydrogen peroxide system resulted in elimination of *C. difficile*, while sodium hypochlorite left 3% of samples still infected. Surface areas sampled included bedside table and washbasin (identified as flat surfaces), handle or inlet of the nightstand drawer, light and nursing-call devices, drip stand, foot bed tubular, inner door-handle of the room, and inner bathroom door handle (identified as irregular surfaces).^22^

The next-generation aerosolized hydrogen peroxide system studied here appears to offer significant efficacy benefits due to droplet dispersion physics and automation. Furthermore, aerosolized hydrogen peroxide drives operational efficiency through the reduction of manual cleaning requirements, delivering rapid cycle times, and removing the logistical need to seal the room. Cloud-based monitoring of activity and recording of results provides additional advantages around compliance and reporting.

Limitations of this study include the fact that the setting was a non-clinical site, with different “obstacles” or shape of room than might be encountered in clinical settings. Although a variety of obstacles were in place in the room, clinical settings may differ. Additionally, air flow in an office setting may vary from that of clinical, operating, or patient care rooms, and general environmental conditions can vary from site to site. This study also evaluated standardized biological indicators rather than naturally contaminated curtains, which may exhibit different bioburden distribution, organic soil burden, and microbial diversity. However, the spores utilized likely represent a conservative biological challenge since spore-forming organisms exhibit substantially greater resistance to disinfection than vegetative bacteria and many enveloped viruses. Finally, this study focused on the objective endpoint of effects on a biological indicator.

## Conclusions

A next-generation aerosolized hydrogen peroxide dry-fogging disinfection system achieved full inactivation (≥4-log_10_ spore reduction) of resistant spores on both sides of privacy curtains within a short 11-minute automated cycle.

## Data Availability

All data produced in the present study are available upon reasonable request to the authors.

